# Somatic structural variation signatures in pediatric brain tumors

**DOI:** 10.1101/2023.05.18.23290139

**Authors:** Yang Yang, Lixing Yang

## Abstract

Brain cancer is the leading cause of cancer-related death in children. Somatic structural variations (SVs), large scale alterations in DNA, remain poorly understood in pediatric brain tumors. Here, we detect a total of 13,199 high confidence somatic SVs in 744 whole-genome-sequenced pediatric brain tumors from Pediatric Brain Tumor Atlas. The somatic SV occurrences have tremendous diversity among the cohort and across different tumor types. We decompose mutational signatures of clustered complex SVs, non-clustered complex SVs, and simple SVs separately to infer the mutational mechanisms of SV formation. Our finding of many tumor types carrying unique sets of SV signatures suggests that distinct molecular mechanisms are active in different tumor types to shape genome instability. The patterns of somatic SV signatures in pediatric brain tumors are substantially different from those in adult cancers. The convergence of multiple signatures to alter several major cancer driver genes suggesting the functional importance of somatic SVs in disease progression.

## Introduction

Brain and central nervous system cancers are the most prevalent solid tumors in children under 19 and are the leading cause of cancer-related death among children (Ostrom et al., 2015). There are more than 100 types of pediatric brain tumors, which differ markedly from adult brain tumors (Louis et al., 2007). Although the 5-year survival rate of pediatric brain tumors is 75%, the survivors often suffer over their lifetimes from both the disease and the side effects of treatments. Therefore, there is an urgent need to better understand the disease mechanisms and to develop new therapeutic strategies in order to further increase survival and improve the quality of life for patients and their families.

Genetic alterations in cancer include single nucleotide variants (SNVs), copy number variants (CNVs), and structural variations (SVs). Pediatric brain tumors have few somatic SNVs, but carry more somatic SVs than other pediatric cancers (Gröbner et al., 2018). SVs are large scale structural changes of DNA such as deletions, tandem duplications, inversions, translocations, and some can be quite complex. For example, chromothripsis refers to a single catastrophic event resulting in numerous SVs through one cell cycle (Maciejowski et al., 2015; Stephens et al., 2011; Zhang et al., 2015). Understanding the mechanisms behind these alterations not only can improve our knowledge of disease biology, but also can reveal therapeutic opportunities. For instance, translocations at the immunoglobulin gene locus in B cell lymphoma are caused by aberrant V(D)J recombination (Alt et al., 2013; Hakim et al., 2012) and often result in activations of *MYC* and *BCL2* oncogenes (Bakhshi et al., 1985; Gostissa et al., 2009). Furthermore, breast and ovarian cancer patients carrying *BRCA1* and *BRCA2* mutations have deficiency in DNA double strand break repair and elevated level of somatic SVs in their tumors (Lee et al., 2016; Nik-Zainal et al., 2016). Patients with BRCA deficiency can be effectively treated by PARP inhibitors (Farmer et al., 2005; Fong et al., 2009). Mutational signatures have been widely used to study the molecular mechanisms of SNVs (Alexandrov et al., 2013, 2020; Degasperi et al., 2022), CNVs (Drews et al., 2022; Steele et al., 2022), SVs (Li et al., 2020) and complex SVs (Bao et al., 2022) in adult cancers. However, comprehensive studies of somatic SV signatures in pediatric brain tumors are still lacking. A recent study of SV signatures in pediatric high-grade glioma has revealed that histone genes, *TP53*, *CDKN2A*, and *RB1* aberrations are associated with complex SVs (Dubois et al., 2022). However, whether other types of pediatric brain tumors harbor similar SV signatures remains unclear.

Here, we decompose complex and simple SV signatures from 744 pediatric brain tumors. We find tremendous heterogeneity in SV occurrences and SV signatures across tumor types. We also report that somatic SVs are frequent cancer drivers in pediatric brain tumors.

## Results and Discussion

### High confidence somatic SVs in pediatric brain tumors

Pediatric Brain Tumor Atlas (PBTA) has collected more than 1,000 pediatric brain tumors across more than 30 types. There were 744 samples in PBTA with whole-genome sequencing data after removing non-tumorous lesions, non-brain cancer, and non-primary cancer samples (Table S1). We focused on tumor types with at least 10 samples including: 220 low-grade astrocytic tumors (LGATs), 97 medulloblastomas, 71 ependymomas, 70 high-grade gliomas (HGGs), 44 gangliogliomas, 38 craniopharyngiomas, 27 atypical teratoid rhabdoid tumors (ATRTs), 23 meningiomas, 23 dysembryoplastic neuroepithelial tumors (DNETs), 17 non-meningothelial mesenchymal tumors, 13 schwannomas, 13 germ cell tumors, 13 neurofibromas, and 12 choroid plexus papillomas. Tumor types with less than 10 samples were all classified as “Others”.

A previous study used Manta (Chen et al., 2016) to detect somatic SVs in the PBTA cohort to study the effects of SVs on gene expression (Zhang et al., 2021). However, the quality of variants called by a single algorithm is always not ideal (Campbell et al., 2020). In order to produce high confidence somatic SVs, we integrated three SV calling algorithms: Meerkat (Yang et al., 2013), Manta and Delly (Rausch et al., 2012a). Caller-specific SVs were discarded, and the somatic SVs detected by more than one algorithm were considered as high confidence. Meerkat, Manta and Delly detected 14,423, 55,934 and 9,475 somatic SVs in the 744 samples, respectively (Figure S1A). Since tumor DNA was not available, the SV quality could not be directly measured. Instead, we used CNV breakpoints detected by read depth approach to assess the quality of SVs, since a portion of somatic SVs change DNA copy numbers. We found SVs detected by only one algorithm were not well supported by CNVs (Figure S1B), which suggested that caller-specific SVs had poorer quality. SVs detected by Manta with fewer read pair and split read support were particularly of poor quality (Figure S1C). SVs detected by more than one algorithm were better supported by CNVs (Figure S1B), which suggested that they were of high quality. We also removed deletions that resided at exon-intron boundaries which were likely caused by cDNA contamination (Campbell et al., 2020). As a result, a total of 13,199 high confidence SVs were detected from 744 pediatric brain tumors with a median of 3 SVs per sample. In each type of pediatric brain tumor, the number of somatic SVs per sample varied by nearly three orders of magnitude (Figure 1A). There was also considerable heterogeneity across tumor types. HGGs were most abundant in somatic SVs followed by meningiomas and medulloblastomas, whereas choroid plexus papillomas had no SV at all (Figure 1A). Although most LGATs, ependymomas, gangliogliomas, and neurofibromas had very few SVs, a small fraction of them had very unstable genomes with more than 100 SVs (Figure 1A).

**Figure 1.**
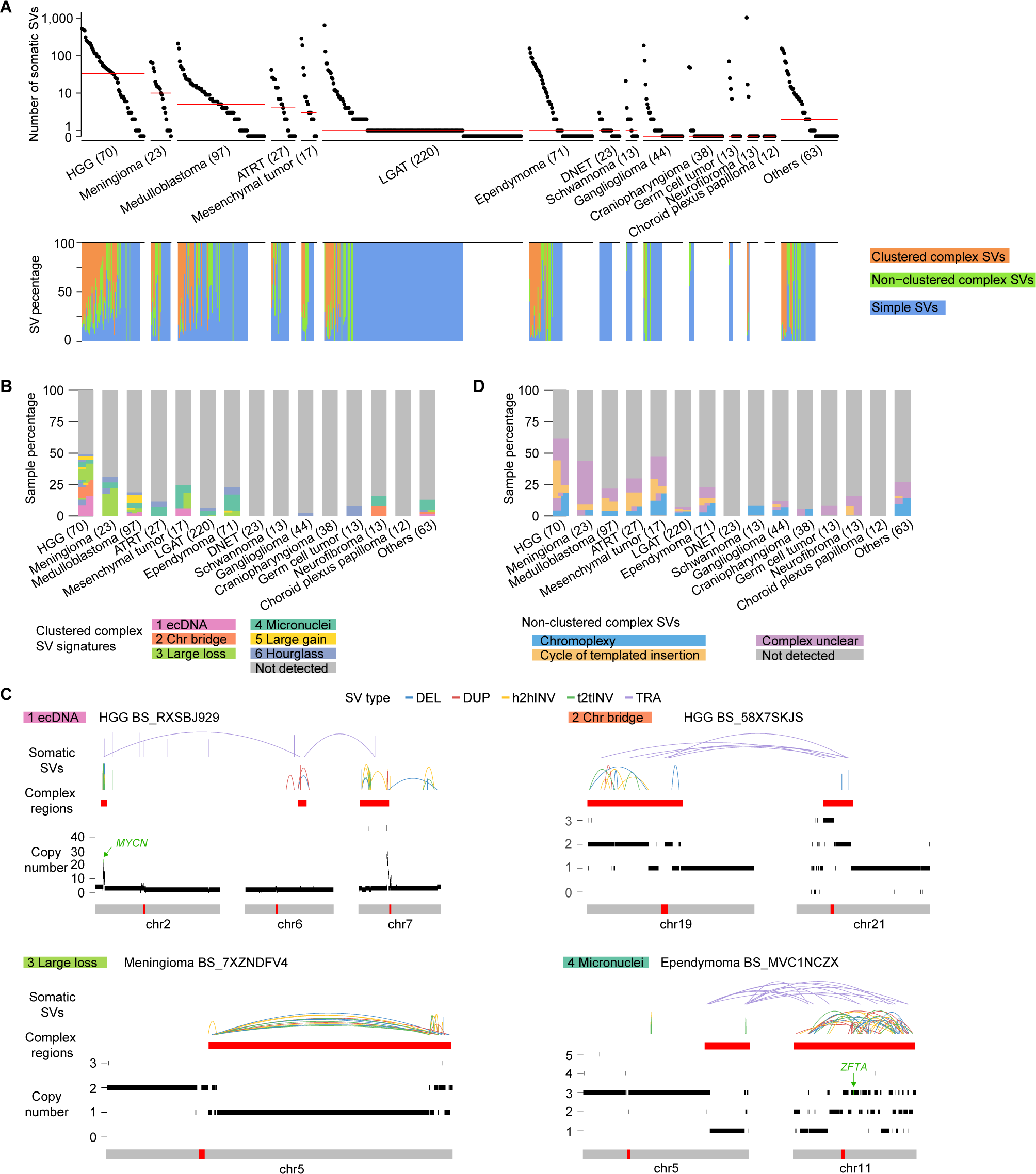
Somatic SVs and complex SVs in 744 pediatric brain tumors. (A) Frequencies of somatic SVs and percentages of different types of SVs. In the upper panel, each dot represents one pediatric brain tumor sample. Samples are grouped by tumor type and tumor types are sorted by median SV frequency (red lines) except for the “Others” category. The numbers in parentheses are sample sizes for the corresponding tumor types. The bottom panel shows the percentages of clustered complex SVs, non-clustered complex SVs, and simple SVs in the corresponding samples on the top panel. HGG: high-grade glioma. ATRT: atypical teratoid/rhabdoid tumor. LGAT: low-grade astrocytic tumor. DNET: dysembryoplastic neuroepithelial tumor. (B) and (D) Percentages of clustered complex SV signatures and non-clustered complex SVs. Each vertical block represents one tumor type and each horizontal bar represent one sample. Samples are colored based on their SV signatures. Samples carry multiple signatures have multiple colors arranged horizontally. The height of each sample may differ across tumor types depending on sample sizes of the tumor types. (C) Examples of clustered complex SVs. Colored arcs represent SVs of different types. The red bars below the SVs indicate regions of clustered complex SVs. Copy number profiles are displayed by black bars above the chromosome models. The red bars within the grey chromosome models indicate the locations of centromeres. Tumor types and sample IDs are shown next to the names of clustered complex SV signatures. DEL: deletion, DUP: tandem duplication, h2hINV: head to head inversion, t2tINV: tail to tail inversion, TRA: translocation.

### Complex SVs in pediatric brain tumors

A non-negative-matrix-factorization-(NMF)-based approach has been very effective in decomposing mutational signatures for somatic SNVs (Alexandrov et al., 2013, 2020; Degasperi et al., 2022) and CNVs (Drews et al., 2022; Steele et al., 2022). Several studies, including the recent SV signature study in pediatric high-grade glioma (Dubois et al., 2022), also used this approach to extract SV signatures by combining both complex SVs and simple SVs (Li et al., 2020). Meaningful signatures can be reliably detected if DNA damage and repair mechanisms generate variants independently and repeatedly in cancer genomes. However, it is well-established that chromothripsis events occur as one-time events and the numbers of SVs vary extensively (Maciejowski et al., 2015; Shoshani et al., 2020; Zhang et al., 2015). Furthermore, multiple molecular mechanisms can lead to chromothripsis. For example, lagging chromosomes trapped in micronuclei during mitosis can shatter into many pieces and some fragments can be ligated together as chromothripsis events (Zhang et al., 2015). Dicentric chromosomes can form chromatin bridges during cell division, shatter into pieces, and also produce chromothripsis (Maciejowski et al., 2015). NMF-based SV signature decomposition cannot differentiate these mechanisms. To better deduce SV signatures in pediatric brain tumors, we studied clustered complex SVs, non-clustered complex SVs and simple SVs separately. Clustered complex SVs are those with breakpoints enriched in certain genomic regions including chromothripsis. Circular extrachromosomal DNA (ecDNA) with many SV breakpoints is also a clustered complex SV (Bao et al., 2022). We recently developed Starfish, a clustering-based approach, to infer clustered complex SV signatures based on their SV and CNV patterns (Bao et al., 2022). We reported six clustered complex SV signatures using nearly 2,500 adult tumors including micronuclei-induced chromothripsis, chromatin-bridge-induced chromothripsis, as well as ecDNA. There are three other signatures that cannot be linked to biological processes, namely “Large loss”, “Large gain” and “Hourglass”. Non-clustered complex SVs are complex SVs with scattered breakpoints including chromoplexy and cycle of templated insertions (Li et al., 2020). Chromoplexy events are likely to form through repair of multiple co-occurring DNA double strand breaks similar to reciprocal translocations (Baca et al., 2013; Berger et al., 2011), whereas templated insertions may reflect replication-based mechanisms (Li et al., 2020; Yang et al., 2013; Zhang et al., 2009). After detecting clustered and non-clustered complex SVs, the remainder of SVs were classified as simple SVs, which include deletions, tandem duplications, balanced/unbalanced/foldback inversions, and balanced/unbalanced translocations.

Among the 13,199 SVs in 744 pediatric brain tumors, 7,601 (57.6%) were clustered complex SVs that belonged to 146 individual complex events, 2,377 (18.0%) were non-clustered complex SVs that belonged to 346 events, and 3,221 (24.4%) were simple SVs (Table S2). Out of the 744 tumors, 108 (14.5%) and 150 (20.2%) carried clustered and non-clustered complex SVs respectively, whereas 552 (74.2%) did not have any complex SVs. The high numbers of SVs in tumors with very unstable genomes (>100 SVs) were mainly due to their abundance of complex SVs (Figure 1A and Figure S2). HGGs were most abundant in complex SVs, whereas DNETs did not carry any complex SVs (Figure 1A). We used Starfish (Bao et al., 2022) to classify clustered complex SV signatures (Table S3) and used junction pattern (Li et al., 2020) to determine non-clustered complex SVs. HGGs and medulloblastomas carried almost all complex SV signatures, whereas other tumor types only harbored small numbers of complex SV signatures (Figure 1B). The “ecDNA” (Figure 1C) signature was predominantly found in HGGs; the “Large loss” signature (Figure 1C), characterized by complex SVs with large amount of DNA loss, was mainly observed in HGGs and meningiomas; micronuclei-induced chromothripsis (“Micronuclei” signature) (Figure 1C) was enriched in mesenchymal tumors and ependymomas (Figure 1B). Chromatin-bridge-induced chromothripsis (“Chr bridge” signature) (Figure 1C) only occurred in HGGs and neurofibromas (Figure 1B). Hourglass chromothripsis events (“Hourglass” signature), complex SVs with a small amount of DNA loss and highly concentrated SV breakpoints, were detected in a small number of samples in several tumor types (Figure 1B), such as HGGs, meningiomas, medulloblastomas, and ependymomas. Regarding to non-clustered complex SVs, chromoplexy was found in many different tumor types and occurred in as many as 18.6% (13/70) of HGGs and 17.6% (3/17) of mesenchymal tumors (Figure 1D). Cycle of templated insertions was abundant in HGGs (Figure 1D).

In summary, different types of pediatric brain tumors often carry distinct complex SV signatures.

### Simple SVs in pediatric brain tumors

Next, we used the NMF approach to decompose simple SV signatures; a total of nine signatures were extracted (Figure 2A and Table S2). These signatures included deletions smaller than 1kb (“Del0”), deletions between 1kb to 5kb (“Del1”), deletions larger than 5kb (“Del2”), tandem duplications (“TD”), unbalanced inversions (“Unbal inv”), large intra-chromosomal SVs (“Large mixed”), reciprocal inversions and reciprocal translocations (“Recip”), as well as unbalanced translocations (“Unbal tra”). Interestingly, large tandem duplications resulting in *KIAA1549*-*BRAF* fusions belonged to a stand-alone signature, namely “*BRAF* fusion”. HGGs had more simple SVs than other tumor types, while a considerable number of samples from various tumor types, including LGATs and ependymomas, showed no evidence of simple SVs (Figure 2B). Most tumors with simple SVs carried multiple simple SV signatures (Figure 2B), and apparent enrichments could be observed. For example, DNETs predominantly carried the “TD” signature; schwannomas mainly harbored the “Recip” signature; the “Unbal inv” signature was mainly found in HGGs; the “TD” signature was enriched in medulloblastomas; and “Del2” signature was abundant in ATRTs; and the “*BRAF* fusion” signature was almost exclusively observed in LGATs (Figure 2B). Intriguingly, 102 out of 220 LGATs had *KIAA1549*-*BRAF* fusions, and among the fusion positive LGATs, 88 had only one SV present in their genomes. The SVs producing *KIAA1549*-*BRAF* fusions were exclusively tandem duplications between 1Mb and 2.5Mb in size (Figure 2A).

**Figure 2.**
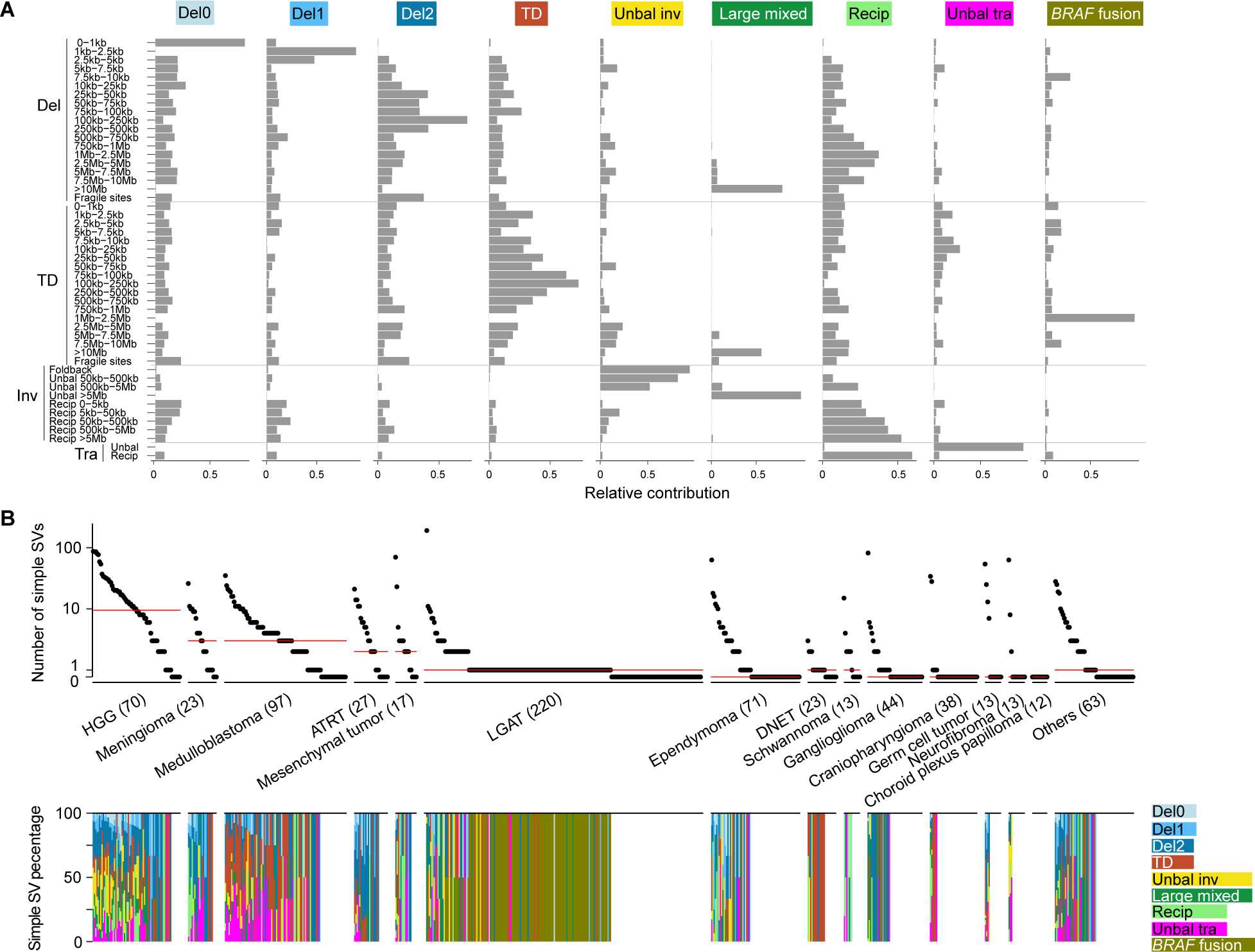
Simple SV signatures and their distributions. (A) Nine simple SV signatures of 744 pediatric brain tumors. The four major SV categories and 49 subcategories of simple SVs are shown on the y axis. The names of the nine simple SV signatures are displayed on the top. The relative contributions of SV subcategories to the corresponding signatures are shown on the x axis. (B) Frequencies of simple SVs and percentages of simple SV signatures. In the upper panel, each dot represents one sample. Samples are grouped by tumor types. Red bars indicate median frequencies. The numbers in parentheses are sample sizes for the corresponding tumor types. The bottom panel shows the percentages of simple SV signatures in the corresponding samples on the top panel.

Taken together, our complex SV and simple SV signature analysis demonstrated that numerous mutational mechanisms are active in pediatric brain tumors to induce genome instability, with unique molecular mechanisms present in different tumor types.

### Genomic features associated with SV signatures

Somatic SVs are not evenly distributed across the genome (Li et al., 2020). Many factors, such as replication timing, GC content, repeat content, and 3D genome organization, have been associated with SV breakpoint distribution (Akdemir et al., 2020; Li et al., 2020). Here, we surveyed 31 genomic features for their relationships with somatic SVs in pediatric brain tumors (Figure 3).

**Figure 3.**
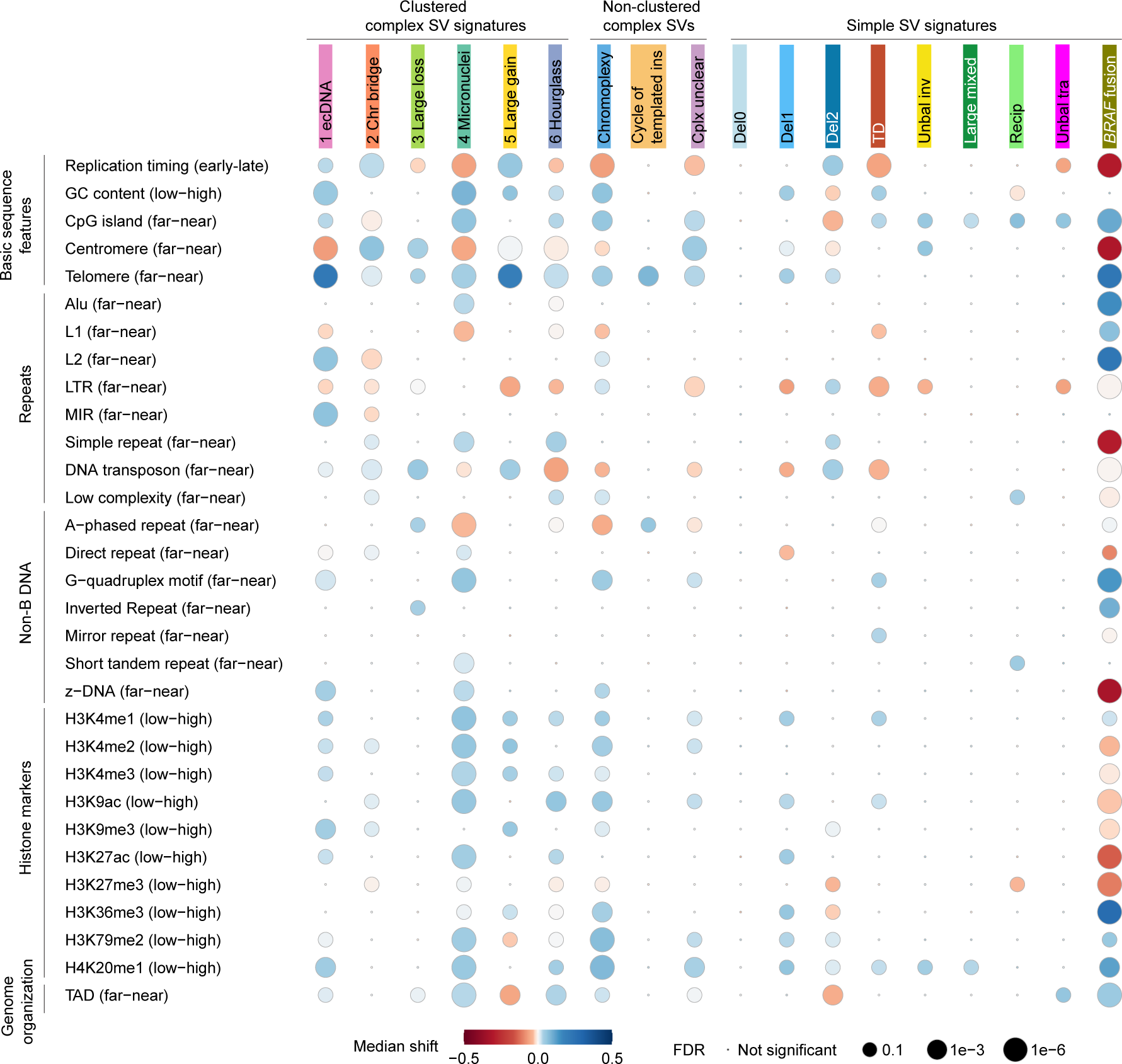
Associations of SV signatures with 31 genomic features. SV signatures and genomic features are listed in the x and y axes respectively. Each dot represents the association between one SV signature and one genomic feature. The bigger the dots are, the more significant the associations are. The colors of the dots indicate the directions of median shift. The directions of biases are shown in the parentheses of genomic features. For instance, the “ecDNA” signature has red color (left on the color key) dot for centromere, indicating that the observed SV breakpoints of this signature are farther away (left in the parentheses) from centromeres than randomized breakpoints.

For clustered complex SV signatures, SV breakpoints of the “ecDNA”, “Chr bridge” and “Large gain” signatures were significantly enriched in late replicated regions (Figure 3). In contrast, all clustered complex SV signatures in adult cancers were enriched in early replicated regions (Bao et al., 2022). In adult cancers, all clustered complex SV signatures were enriched in GC-rich regions and near CpG islands (Bao et al., 2022), whereas only the “ecDNA” and “Micronuclei” signatures in pediatric brain tumors were enriched in GC-rich regions (Figure 3). SV breakpoints of the “Large loss” signature were significantly closer to centromeres than expected in pediatric brain tumors (Figure 3), in a pattern opposite to adult cancers (Bao et al., 2022). In adult cancers, the “ecDNA” and “Chr bridge” signatures were significantly farther away from telomeres, whereas other clustered complex SVs were significantly closer to telomeres (Bao et al., 2022). However, all clustered complex SV signatures were significantly closer to telomeres in pediatric brain tumors (Figure 3). All clustered complex SV signatures in adult cancers were significantly closer to many types of repetitive elements (simple repeats, short tandem repeats and transposable elements) (Bao et al., 2022), whereas in pediatric brain tumors, the repetitive elements had variable effects. For example, the “Micronuclei” signature in pediatric brain tumors were enriched near Alu elements but depleted around L1s (Figure 3). Therefore, the SV breakpoint distributions of clustered complex SVs in pediatric brain tumors were quite different from those of adult cancers.

Chromoplexy was proposed to form similar to reciprocal translocations through simultaneous ligation of multiple broken chromosomal ends (Baca et al., 2013; Berger et al., 2011). In adult cancers, chromoplexy breakpoints and reciprocal translocations shared similar patterns. For example, they were both enriched in late replicated regions. In sharp contrast, chromoplexy breakpoints in pediatric brain tumors were enriched in early replicated regions while reciprocal translocations did not display any bias in replication timing (Figure 3). In addition, chromoplexy breakpoints in pediatric brain tumors were also enriched in GC-rich regions and near telomeres, while reciprocal translocations were enriched in AT-rich regions and not enriched towards either centromeres or telomeres (Figure 3). These results suggested that chromoplexy in pediatric brain tumors may form through a different mechanism than reciprocal translocations. Cycle of templated insertion breakpoints in pediatric brain tumors had little association with most genomic features except for proximity to telomeres (Figure 3).

Simple SV breakpoint distributions of pediatric brain tumors were also quite distinct from those of adult cancers. In adult cancers, deletions were enriched in early replicated regions, and tandem duplications and unbalanced translocations were enriched in late replicated regions (Li et al., 2020). In contrast, in pediatric brain tumors, smaller deletions (“Del0” and “Del1”) were not associated with replication timing, large deletions (“Del2”) were enriched in late replicated regions, and tandem duplications (“TD”) as well as unbalanced translocations (“Unbal tra”) were enriched in early replicated regions (Figure 3). In adult cancers, all deletions regardless of their sizes were enriched in AT-rich regions (Li et al., 2020); whereas in pediatric brain tumors, small deletions (“Del1”) were enriched in GC-rich regions and large deletions (“Del2”) were enriched in AT-rich regions (Figure 3). In adult cancers, tandem duplications were significantly closer to Alu elements (Li et al., 2020); whereas in pediatric brain tumors, tandem duplications (“TD”) were farther away from L1s, LTR transposons, and DNA transposons (Figure 3). In adult cancers, both small and large deletions were depleted from topologically associated domain (TAD) boundaries (Li et al., 2020). However, in pediatric brain tumors, only large deletions (“Del2”), but not small deletions (“Del0” and “Del1”), were depleted from TAD boundaries (Figure 3).

Taking the above results together, our results suggest that the forming mechanisms of somatic SVs, both complex and simple SVs, in pediatric brain tumors were likely to be different from those of adult cancers.

Next, we sought to identify mutations associated with genome instability. As expected, *TP53* mutations in HGGs were associated with “Chr bridge” and “Large loss”, and cycle of templated insertions, as well as six simple SV signatures: “Unbal inv”, “Unbal tra”, “Large mixed”, “TD”, “Del1” and “Del2” (Figure S3). In addition, *H3F3A* mutations in HGGs were associated with cycle of templated insertions (Figure S3) suggesting that histones may play important roles in genome instability in pediatric brain tumors. *ATRX* mutations in HGGs were associated with “Large mixed” signatures (Figure S3). No other mutations were associated with any SV signatures in any other pediatric brain tumors. In adult cancers, small deletions are associated with *BRCA2* mutations, and small and large tandem duplications are associated with *BRCA1* and *CDK12* mutations (Li et al., 2020; Nik-Zainal et al., 2016). In the 741 non-hypermutated pediatric brain tumors, only one sample carried a *BRCA1* mutation, and another sample carried a *BRCA2* mutation. This again suggested that the mechanisms of formation of deletions and tandem duplications in pediatric brain tumors are likely to be different from those of adult cancers.

### SV breakpoint sequences

We then investigated microhomology and insertion sequences at the SV breakpoints across SV signatures. SVs are the results of erroneous repair of DNA double strand breaks or replication errors. Various repair pathways are involved (Aguilera and García-Muse, 2013; Bunting and Nussenzweig, 2013), such as non-homologous end joining (NHEJ), alternative end joining (alt-EJ), and microhomology-mediated break-induced repair (MMBIR). NHEJ usually ligates blunt DNA ends or ends with short 1-4bp homology. Alt-EJ often uses slightly longer homology for repair. MMBIR is considered a replication based template switching mechanism and non-template insertions are frequently present at the breakpoints (Zhang et al., 2009). Several SV signatures in pediatric brain tumors, including “Large loss”, “Hourglass”, chromoplexy, and “Unbal tra”, had a majority of SV breakpoints being blunt DNA ends (no homology nor insertion at the breakpoints) (Figure 4). This suggested that these SVs are likely to form through NHEJ. Some other signatures, such as “Chr bridge”, “Micronuclei”, “Del0”, “Del1”, “Unbal inv”, and “Large mixed”, had slightly longer homology at the breakpoints with 1bp microhomology being the most frequent and also consistent with NHEJ (Figure 4). In addition, “TD”, “Recip”, and “*BRAF* fusion” signatures had 2bp microhomology being the most frequent (Figure 4) suggesting alt-EJ might play more important roles in these SVs. The observation of extended microhomology in the “Recip” signature was consistent with breakpoints found in the Philadelphia chromosome, the most prevalent reciprocal translocation in leukemia, often involving 2-8 bp microhomology (McVey and Lee, 2008). Although previous studies proposed that chromoplexy forms in a manner similar to reciprocal translocation (Baca et al., 2013; Berger et al., 2011), our observation of chromoplexy and reciprocal translocation breakpoints with different microhomology patterns suggested that they may form via different mechanisms. Furthermore, a fraction of SVs in “Del1” had 5 bp or longer microhomology (Figure 4) especially in ependymomas, suggesting alt-EJ being the dominant mechanism in ependymoma. Intriguingly, “ecDNA” and “Large gain” signatures had frequent inserted sequences that were more than 10 bp (Figure 4), suggesting possible involvement of replication-based mechanisms such as MMBIR.

**Figure 4.**
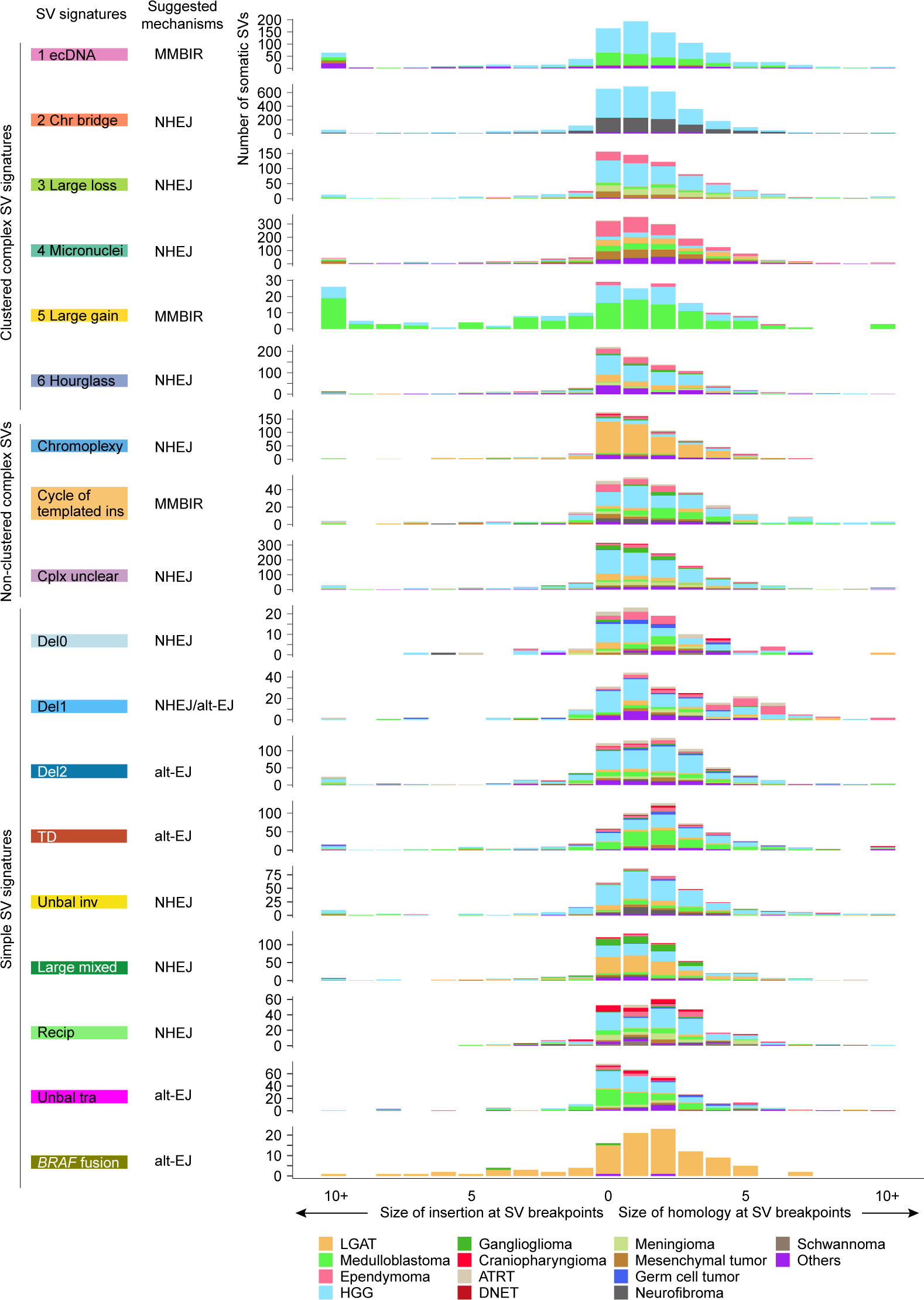
SV breakpoint homology. The distributions (x axis) of homology and insertion at SV breakpoints are shown for all SV signatures (y axis). The putative DNA repair mechanisms are inferred from the sizes of homology and insertion and annotated next to the signatures. The bars indicate number of somatic SVs and are colored by tumor type. MMBIR: microhomology-mediated break-induced repair. NHEJ: non-homologous end joining. alt-EJ: alternative end joining.

### SV hotspots and tumor drivers

Hotspots of SV breakpoints often represent genetic alterations under positive selection and genes driving diseases. After binning the reference genome into 1 Mb window and counting SV occurrences, we found different SV signatures having quite distinct hotspots. *MYCN* and *MYC* are two frequently amplified oncogenes in pediatric brain tumors (Dubois et al., 2022). Interestingly, *MYCN* was amplified exclusively by “ecDNA” in HGGs and exclusively by “Large gain” in medulloblastomas (Figure 5), since “ecDNA” and “Large gain” were the most abundant clustered complex SV signatures in HGGs and medulloblastomas, respectively. Both signatures converged on *MYCN* amplifications, which are the main oncogenic events in HGGs and medulloblastomas. *MYCN* was also amplified by non-clustered complex SV with unclear pattern (“Complex unclear”) in various tumor types (Figure 5). Tandem duplications can also amplify DNA. However, *MYCN* was not amplified by the “TD” signature in any samples, whereas *MYC* was amplified by the “TD” signature in a few HGGs and medulloblastomas (Figure 5). In addition, *FGFR1* was frequently amplified by the “TD” signature in DNETs (Figure 5). DNETs had very few somatic SVs and did not carry any clustered or non-clustered complex SVs. “TD” was the dominant SV signature in DNETs. These findings suggested that “TD” and *FGFR1* amplifications were the major oncogenic events in DNETs. Furthermore, chromoplexy and “Del2” frequently disrupted *CDKN2A* in various tumor types (Figure 5). The “*BRAF* fusion” signature produced *KIAA1549*-*BRAF* fusions mainly in LGATs (Figure 5). The “Recip” signature often led to *EWSR1* fusions in various tumor types (Figure 5).

**Figure 5.**
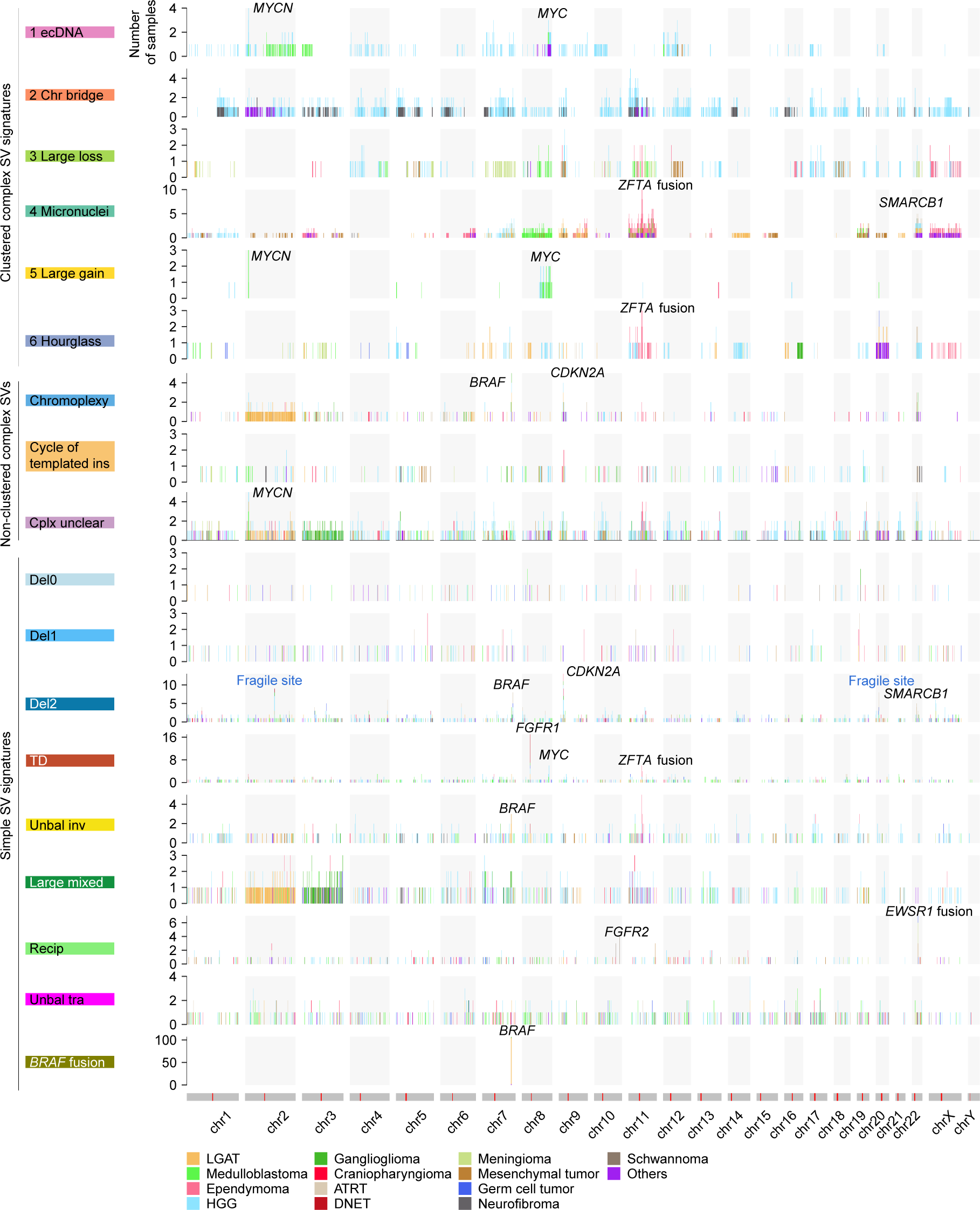
SV breakpoint hotspots. SV breakpoint frequencies are shown for the entire reference genome (x axis) across different SV signatures (y axis). Chromosome models are shown as grey bars with red lines indicating locations of centromeres at the bottom. Hotspots containing known oncogenes, tumor suppressors, and fragile sites are annotated.

Interestingly, multiple SV signatures harbored hotspots on chromosome 11, and the hotspots were only found in ependymomas (Figure 5). *C11orf95*-*RELA* fusions are the major oncogenic events in 70% of supratentorial ependymomas (Parker et al., 2014). Recently, the World Health Organization (WHO) recommended the use of the *ZFTA* (*C110rf95*) fusion to classify supratentorial ependymoma instead of the *RELA* fusion because *ZFTA* can fuse to other partners as well (Louis et al., 2021). There were 71 ependymomas in our cohort and 23 of them (32%) were supratentorial (Figure 6A and Table S4). Among them, 13 (57%) carried *ZFTA* fusions (Figure 6A). There were 2 other *ZFTA* fusion positive ependymomas classified as “Others” (Figure 6A). Twelve out of 15 *ZFTA* fusions were driven by complex SVs and three were driven by tandem duplications (“TD”) (Figure 6A). Among the 12 fusions resulting from complex events, 5 were micronuclei-induced chromothripsis (“Micronuclei”), 3 were hourglass chromothripsis (“Hourglass”), and 4 were non-clustered complex SVs (Figure 6A and 6B). Some of the complex SVs involved the entire chromosome 11 (BS_K6A9Z04J), whereas others only affected a small region in chromosome 11 (BS_NWYBD9CA) (Figure 6B). These results showed that there are diverse mechanisms generating genome instability in ependymomas and they have shared oncogenic consequence of forming *ZFTA* fusions. The most prevalent complex SVs in ependymomas were micronuclei-induced chromothripsis events caused by erroneous chromosomal segregation. It is possible that the frequent complex SVs in ependymomas are due to frequent chromosomal segregation errors. However, aneuploidy is rare in ependymomas (Mack and Taylor, 2017). This suggested that chromosomal segregation errors are likely not frequent events in ependymomas, and complex SVs in ependymomas are under positive selection. Furthermore, since tandem duplications are sufficient to produce gene fusions, such as *ZFTA* fusions and *BRAF* fusions, and the fact that most somatic SVs in ependymomas involving chromosome 11 were complex SVs suggested that other genes altered by SVs may contribute to ependymoma tumorigenesis as well. “Unbal inv” signature also had a hotspot in a similar region on chromosome 11 (Figure 5). Three ependymomas had unbalanced inversions in gene *MARK2*, which is next to *ZFTA*, and no fusions were formed. Whether this “Unbal inv” hotspot in ependymoma reflects oncogenic events remains unclear.

**Figure 6.**
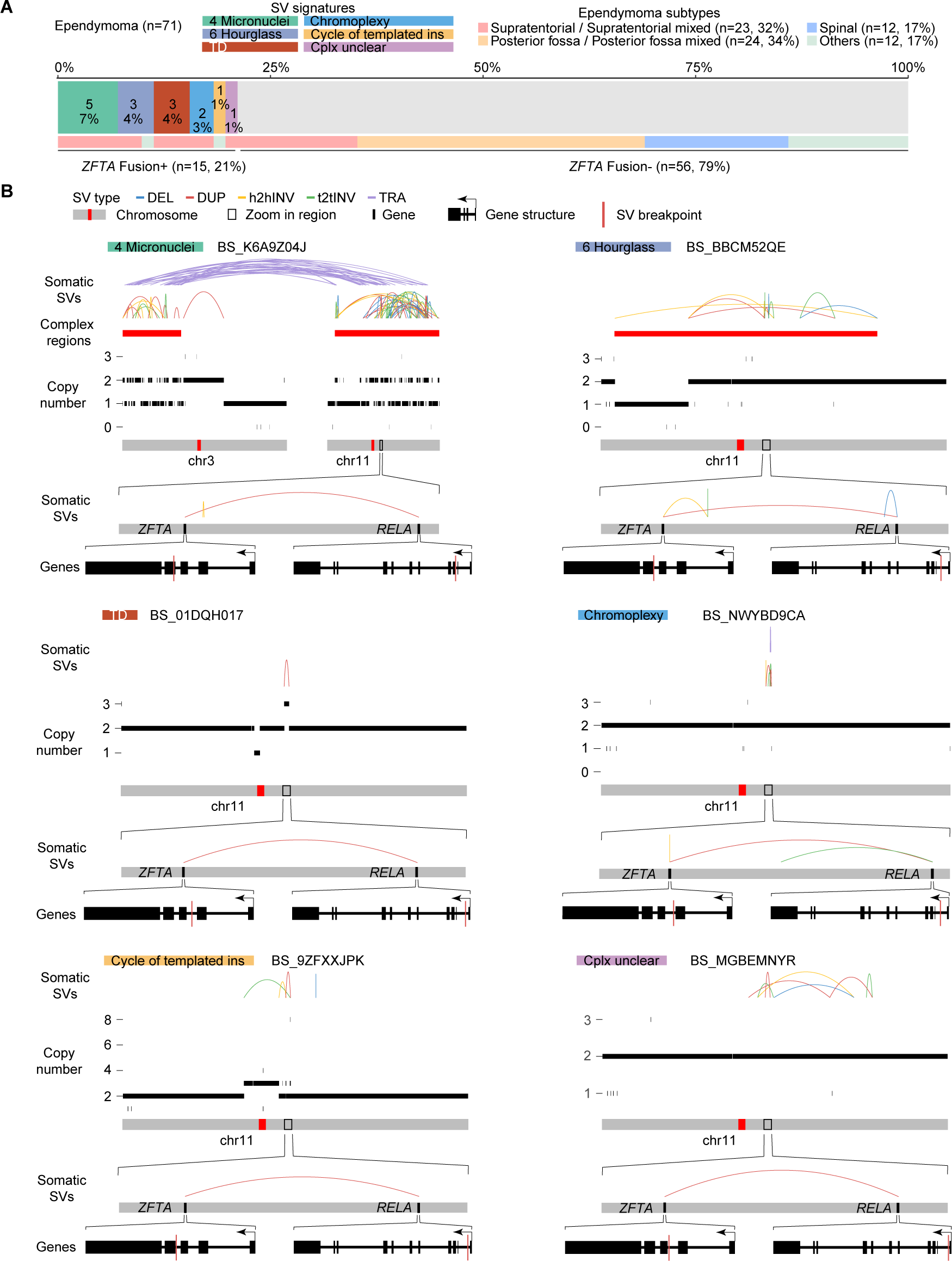
*ZFTA* fusions in ependymomas. (A) The prevalence of *ZFTA* fusions in 71 ependymomas. Samples are colored by signatures of the SVs resulting in *ZFTA* fusions and ependymoma subtypes. (B) Six examples of *ZFTA* fusions resulting from different SV signatures. SV signatures and sample IDs are shown on the top. Somatic SVs, regions of complex SVs and copy number profiles are displayed in the same scheme as Figure 1C. The *ZFTA* and *RELA* regions are zoomed in and *ZFTA* gene and *RELA* gene are further zoomed in respectively. Gene structures are shown at the bottom of six examples. Within gene structures, the SV breakpoints that lead to *ZFTA*-*RELA* fusions are shown as red vertical lines. The directions of gene transcription are indicated by arrows.

In a significant fraction of pediatric brain tumors, the disease-causing SVs were the sole SVs of the corresponding signatures. These signatures did not produce additional passenger SVs in those samples. For example, 88 LGATs were driven by *BRAF* fusions and there was only one SV within the “*BRAF* fusion” signature in those samples (Figure 2B). Similarly, three ependymomas carried *ZFTA* fusions caused by the “TD” signature, and there was only one SV within the “TD” signature in two of these three samples (Figure 2B and Figure 5). In addition, nine DNETs had *FGFR1* amplifications resulting from the “TD” signature, and there was only one SV within the “TD” signature in all nine samples (Figure 2B and Figure 5). These results indicated that the molecular mechanisms leading to these disease-driving SVs are not highly active in tumor initiating cells. Cells independently acquire SVs through these mechanisms at very low rates, and cells that have acquired SVs that alter the major disease-driving genes eventually outcompete other cells and become tumors.

### Impact of SVs on patient survival

Next, we sought to evaluate whether somatic SVs affect patient survival in pediatric brain tumors. Chromothripsis has been associated with worse patient survival in several previous studies (Cortés-Ciriano et al., 2020; Molenaar et al., 2012; Notta et al., 2016; Rausch et al., 2012b). However, we did not observe any complex SV signatures associated with patient survival in HGGs, LGATs, medulloblastomas or ependymomas (Figure S4). HGG patients with “Del2” and “Unbal tra” signatures in their tumors had significantly worse survival, and those with “Del1” and “Large mixed” signatures had marginally worse survival (Figure 7). HGGs were most abundant in simple SVs, which suggested that the SV forming mechanisms are relatively more active in tumor initiating cells of HGGs than those of other tumor types. No simple SV signatures were associated with patient survival in other tumor types. It is possible that simple SV forming mechanisms are not very active in tumor types other than HGGs; therefore simple SV signatures are associated with patient survival only in HGGs.

**Figure 7.**
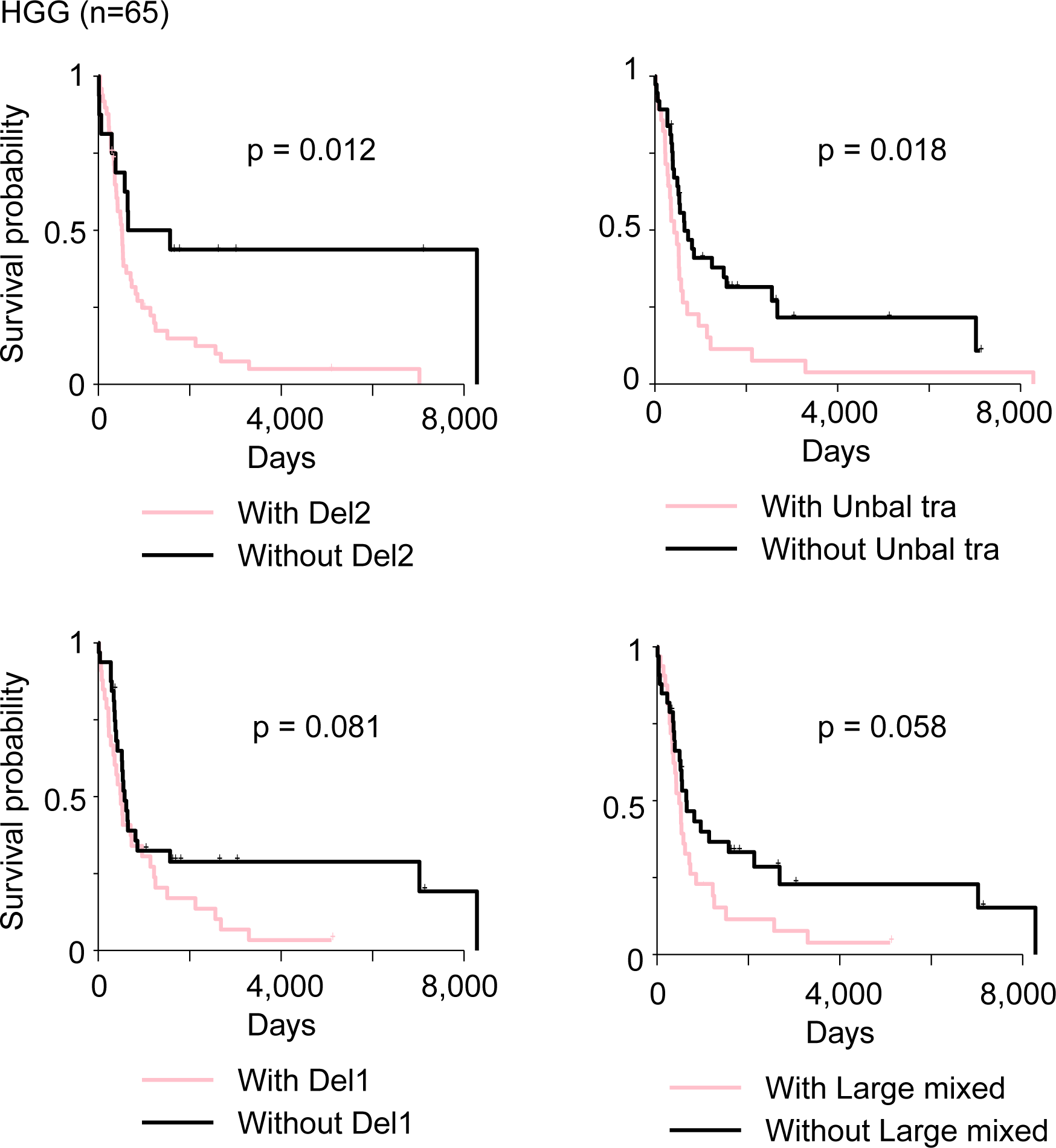
SV signatures associated with patient survival. Kaplan-Meier survival curves for HGG patients, stratified by the presence or absence of four simple SV signatures (“Del2”, “Unbal tra”, “Del1”, and “Large mixed”) are shown. P values are calculated by log-rank test.

## Data Availability

All data produced in the present work are contained in the manuscript

## Acknowledgement

We thank the Center for Research Informatics at the University of Chicago for providing the computing infrastructure. We thank Dr. Ji Won Lee for assistance in tumor pathology. The work was supported by the National Institutes of Health grant R03CA246228 (L.Y.).

## Author contributions

L.Y. conceptualized the study. Y.Y. developed the analysis workflow and performed the analysis. Y.Y. and L.Y. interpreted the results. L.Y. supervised the study. L.Y. wrote the paper. All authors have read and approved the final manuscript.

## STAR Methods

### Sample and data collection

The raw normal and tumor whole-genome sequencing data for 744 pediatric brain tumor patients were downloaded from CAVATICA (https://cavatica.sbgenomics.com/). Sample characteristics, clinical data, somatic SNV, and somatic CNV data were retrieved from OpenPBTA GitHub (https://github.com/AlexsLemonade/OpenPBTA-analysis, https://www.biorxiv.org/content/10.1101/2022.09.13.507832v1). All patients were de-identified.

Gene annotation was obtained from ENSEMBL (GRCh38.p13) (https://useast.ensembl.org/index.html). Non-B DNA structures including A-phased repeats, direct repeats, G-quadruplex forming repeats, inverted repeats, mirror repeats, short tandem repeats and Z-DNA motifs were downloaded from non-B DB (Cer et al., 2013) (https://nonb-abcc.ncifcrf.gov/apps/nBMST/default/); Alu, L1, L2, LTR, MIR, simple repeat, transposon, and low complexity repetitive elements, as well as the coordinates of centromeres, telomeres, and CpG islands, were obtained from UCSC (https://hgdownload.soe.ucsc.edu/goldenPath/hg38/database/); consensus estimate of the topologically associated domains (TADs) were downloaded from TAD Map (Singh and Berger, 2021) (https://cb.csail.mit.edu/cb/tadmap/); ChIP-seq data of epigenetic markers H3K4me1, H3K9me3, H3K36me3, H3K4me3, H3K27me3, H3K9ac, H3K4me2, H3K79me2, H3K27ac, and H4K20me1 from human astrocyte were downloaded from ENCODE (Zhang et al., 2020) (https://www.encodeproject.org/). The Wavelet-smoothed signal of replication timing data for the cell lines BG02ES and SK-N-SH were downloaded from UCSC (https://genome.ucsc.edu/cgi-bin/hgFileUi?db=hg19&g=wgEncodeUwRepliSeq). The fragile site regions were obtained from a previous study (Li et al., 2020), and the coordinates were lifted over to hg38. All coordinates in this study were based on the hg38 genome assembly unless otherwise noted.

### Tumor classifications

The histological classifications of tumor samples were determined based on diagnosis, pathological examination, and histological examination according to the 2021 WHO classifications of pediatric brain tumors (Louis et al., 2021). For tumor types with at least 10 samples, we attempted to subclassify them. Gliomas were subclassified into high-grade gliomas (HGGs), low-grade astrocytic tumors (LGATs), ependymomas, dysembryoplastic neuroepithelial tumors (DNETs), and gangliogliomas. Embryonal tumors were subclassified into medulloblastomas, and atypical teratoid rhabdoid tumors (ATRTs). Cranial and paraspinal nerve tumors included schwannomas and neurofibromas. Germ cell tumors included teratomas, germinomas, and other germ cell tumors. Mesenchymal non-meningothelial tumors included hemangioblastomas, Ewing sarcomas, rhabdomyosarcomas, myofibroblastomas, and other sarcomas. Meningiomas, craniopharyngiomas, and choroid plexus tumors were independently classified. Tumor types with less than 10 samples were all classified into “Others”. Ependymomas were further stratified into distinct subtypes based on the primary sites.

### Somatic SV calling and filtering

Manta (https://github.com/Illumina/manta), Meerkat (https://github.com/guru-yang/Meerkat), and Delly (https://github.com/dellytools/delly) were used for somatic SV detection. SVs called by Manta were obtained from OpenPBTA. Meerkat was run as suggested (Yang et al., 2013). Delly was run with default settings. For SVs detected by Delly, at least four supporting read pairs and split read combined were required for SVs less than 500 bp. For all three SV detection algorithms, only SVs located in canonical chromosomes (chr1-22, X, Y) were retained. SVs identified by different algorithms were considered identical if their two breakpoints were on the same chromosomes, with the same orientations and within 10 bp. SVs identified by two or more algorithms were considered high-confidence SVs and used in the subsequent analysis. Deletions with both breakpoints within 3 bp of exon-intron boundaries of the same genes were excluded from further analysis.

Somatic CNVs were used to assess the quality of somatic SVs. For each SV, if the distances of both SV breakpoints were less than 1 kb to the nearest CNV breakpoints, the SV was considered validated.

### Complex SVs and their signatures

We used Starfish (Bao et al., 2022) (https://github.com/yanglab-computationalgenomics/Starfish) to detect clustered complex SVs and classified them into six signatures. In cases where reported gender and germline estimated sex were inconsistent, gender identity was recorded as unknown for signature detection. After removing clustered complex SVs, we used ClusterSV (Li et al., 2020) (https://github.com/cancerit/ClusterSV) to identify non-clustered complex SVs. Non-clustered complex SVs include chromoplexy, cycle of templated insertions, and complex unclear.

### Simple SV signatures

After removing clustered and non-clustered complex SVs, the remainder were simple SVs including four major categories: deletions, tandem duplications, inversions, and translocations. Deletions and tandem duplications with breakpoints falling within fragile site regions were classified as fragile site deletions and fragile site tandem duplications, respectively. The remaining deletions and tandem duplications were classified into 18 subcategories based on their sizes. Inversions and translocations were further subclassified into reciprocal inversions, fold-back inversions, unbalanced inversions, reciprocal translocations, and unbalanced translocations. Unbalanced inversions and reciprocal inversions were classified into 3 and 5 subcategories based on their sizes, respectively. Fold-back inversions, unbalanced translocations, and reciprocal translocations were three independent subcategories. As a result, all simple SVs were classified into 49 subcategories and SigProfilerExtractor (Islam et al., 2022) (https://github.com/AlexandrovLab/SigProfilerExtractor) with default parameters was used to extract simple SV signatures. According to the final signatures we chose, deletions smaller than 1kb were assigned as “Del0”; deletions ranging in size from 1kb-5kb were classified as “Del1”; fragile site deletions, fragile site tandem duplications, and deletions sized in 5kb-10Mb were assigned as “Del2”; tandem duplications between 1Mb-2.5Mb with breakpoints located within the *BRAF* region were classified as *BRAF* fusion signature; other tandem duplications were classified as “TD”; foldback inversions and unbalanced inversions sized between 50kb-5Mb were categorized as “Unbal inv”; deletions and tandem duplications larger than 10Mb, as well as reciprocal unbalanced inversions larger than 5Mb, were classified as “Large mixed”; reciprocal inversions and reciprocal translocations were categorized as “Recip”; and unbalanced translocations were classified as “Unbal tra”.

### Genomic feature tests

For each observed somatic SV, we generated four random SVs on the same chromosome, same size, and same type. All observed and randomized breakpoints were annotated with genomic features. Bedtools was used to compute the GC content within a ±50 bp interval of each SV breakpoint. The distances in kilobases (kb) from the breakpoints to the nearest Non-B DNA structures, repetitive elements, and CpG islands were logarithmically transformed, with the distances set to 0 if breakpoints were within any of the aforementioned elements. The distances in megabases (Mb) from the breakpoints to centromeres and telomeres and the distances (kb) to the closest TAD boundaries were also transformed to log scale. The SV breakpoints were annotated by signal -log10(p-values) for different epigenetic modifications. The replication timing data for cell lines BG02ES and SK-N-SH were quantile normalized. The SV breakpoints were lifted over to hg19 since the replication timing data were based on hg19. The replication timing values were then annotated for each SV breakpoint. Breakpoints of observed SVs and randomized SVs were tested as described in the previous study (Li et al., 2020). Briefly, scores of SV breakpoints for all genomic features were rescaled from 0 to 1. The distributions of scores between observed breakpoints and randomized breakpoints were compared using two-sided Kolmogorov-Smirnov test. False discovery rates (FDRs) were computed using the Benjamini-Hochberg procedure and 0.1 FDR cutoff was used to determine significant associations. Homology and insertion size at the SV breakpoints were provided by Meerkat and Manta.

### Hotspot analysis

The reference genome was divided into 1Mb non-overlapping bins. The number of samples with SV breakpoints in each bin was counted for each SV signature. A sample with multiple SV breakpoints of the same SV signatures falling within the same bin was only counted once.

### Mutation test

Only protein-altering somatic SNVs and indels were considered, including missense mutations, splice site mutations, frameshift indels, nonsense mutations, translation start site mutations, and nonstop mutations. Three HGG samples (BS_20TBZG09, BS_02YBZSBY, and BS_VW4XN9Y7) with hypermutation were excluded. The tests were performed within tumor types. Protein-coding genes with mutation frequencies >= 5% in each tumor type were analyzed. Samples were classified into two categories based on the presence and absence of the SV signatures. Fisher’s exact test was used to calculate p values. FDRs were computed using the Benjamini-Hochberg procedure. FDR < 0.1 was considered as significant.

### Survival analysis

Since patient survival differs dramatically across tumor types, survival analysis was only performed within tumor types, but not across tumor types. For clustered complex SV signatures, samples with only one signature were assigned to the corresponding signatures; samples with more than one clustered complex SV signatures were classified into “Mixed”; and samples without any clustered complex SV were assigned into “None”. For simple SV signatures, samples were classified based on the presence and absence of the signatures. Log-rank test was used to calculate p values.

