## Supplementary Figures for "Somatic structural variation signatures in pediatric brain tumors"

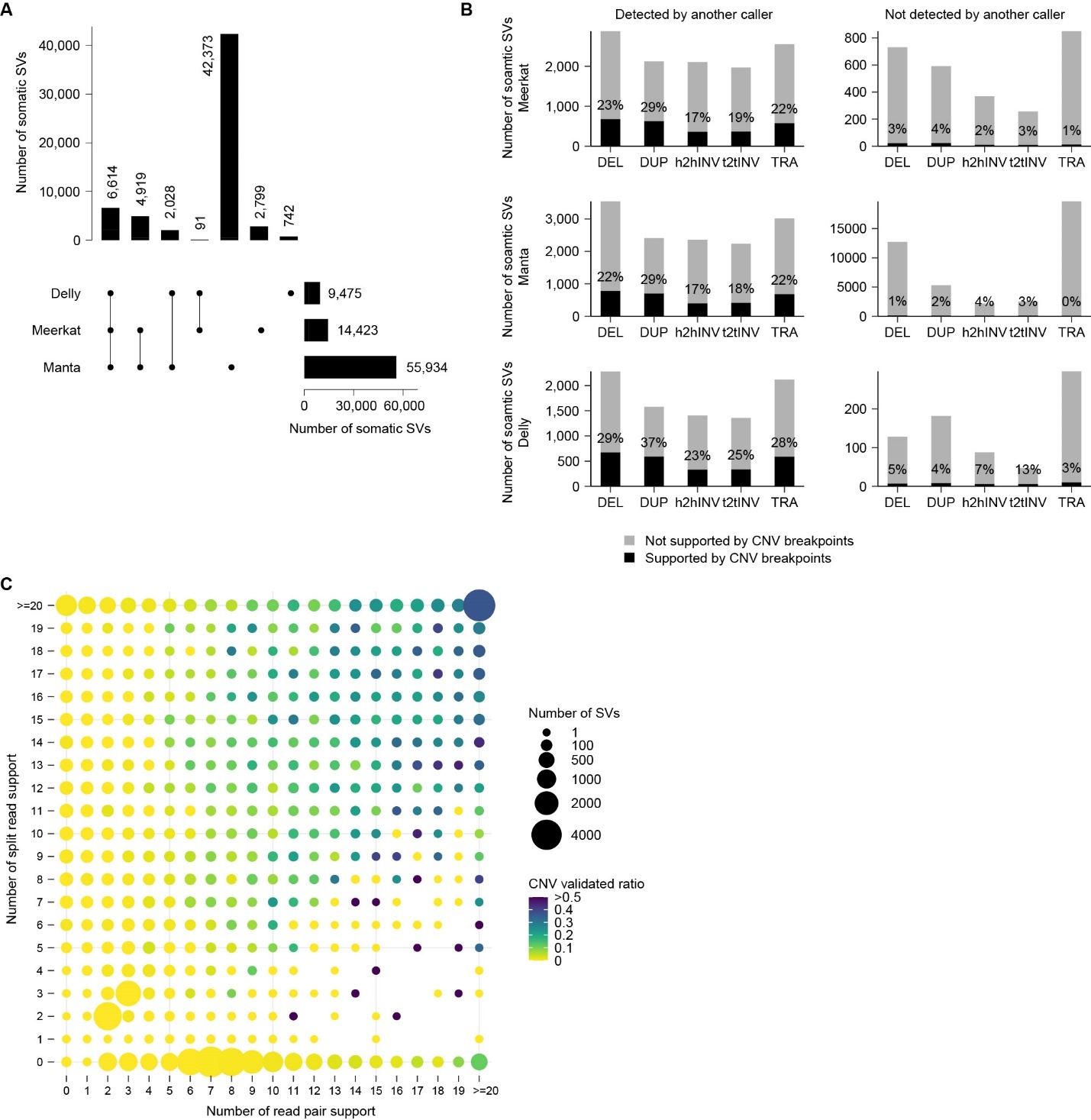
 **Figure S1 Quality control of somatic SVs called by Manta, Meerkat, and Delly.** (A) UpSet plot of somatic SVs called by Meerkat, Manta, and Delly. The bars on the bottom right are the total SVs detected by the three tools respectively. The bars on the top show the number of SVs identified by one or more tools. The black dots under the bars indicate tools used. The numbers on the top and on the right side of the bars are numbers of SVs. (B) SVs called by different tools validated by CNVs. The left panels are high confidence SVs called by more than one tools, and the right panels are tool-specific low confidence SVs. The black and grey bars indicate SVs supported or not supported by CNV breakpoints, respectively. (C) CNV validation of Manta SVs. The x and y axes are the number of read pair and number of split read supports for SVs detected by Manta, respectively. The size of each dot represents the number of SVs, and the color indicates the proportion of SVs validated by CNVs.

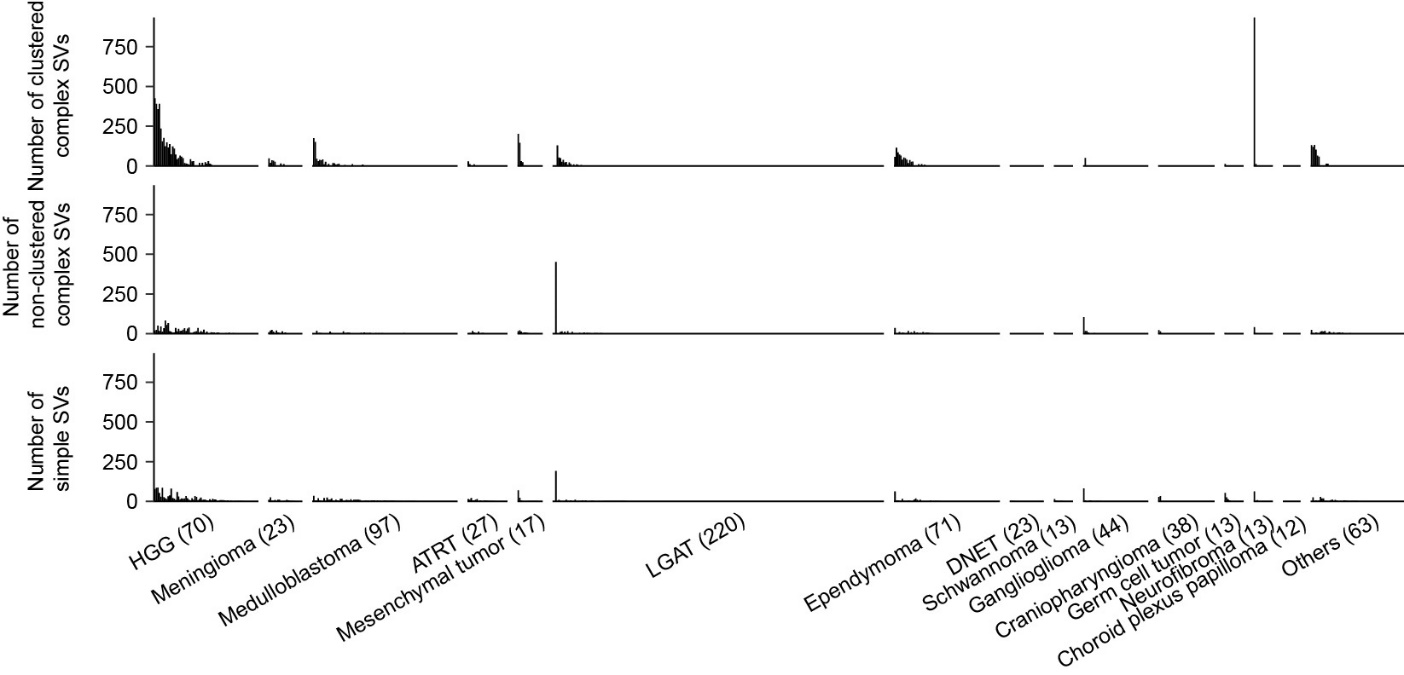

**Figure S2 Frequencies of complex SVs and simple SVs.** The three tracks present the number of clustered complex SVs, non-clustered complex SVs, and simple SVs, respectively. The tumor type ordering is consistent with Figure 1, and samples are sorted by the total number of SVs. The numbers in parentheses represent the sample sizes for the corresponding tumor types.

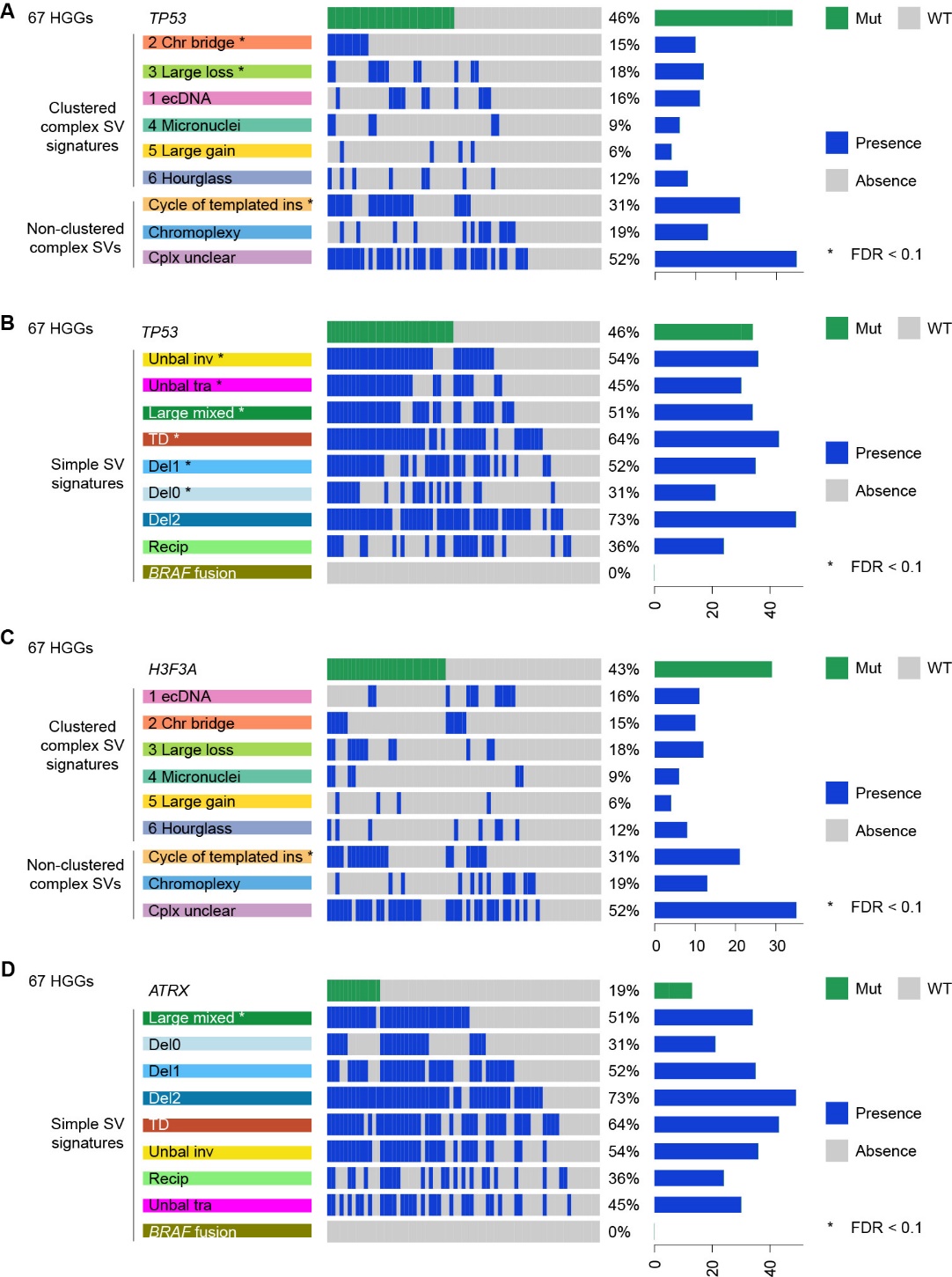

**Figure S3 Associations between SV signatures and somatic mutations in protein-coding genes in 67 non-hypermutated HGGs.** (A) The associations between *TP53* mutations and complex SV signatures in HGGs. (B) The associations between *TP53* mutations and simple SV signatures in HGGs. (C) The associations between *H3F3A* mutations and complex SV signatures in HGGs. (D) The associations between *ATRX* mutations and simple SV signatures in HGGs. Protein-altering mutations are represented by green bars, while the presence of signatures is indicated by blue bars. The percentages reflect the proportions of samples carrying the corresponding mutations or signatures. The horizontal bars represent the frequencies of the corresponding mutations or signatures in HGGs. The associations were assessed using Fisher's exact test, and the FDRs were adjusted using the Benjamini-Yekutieli method. An asterisk (*) denotes a statistically significant positive correlation (FDR < 0.1) between the tested genes and the corresponding signatures.

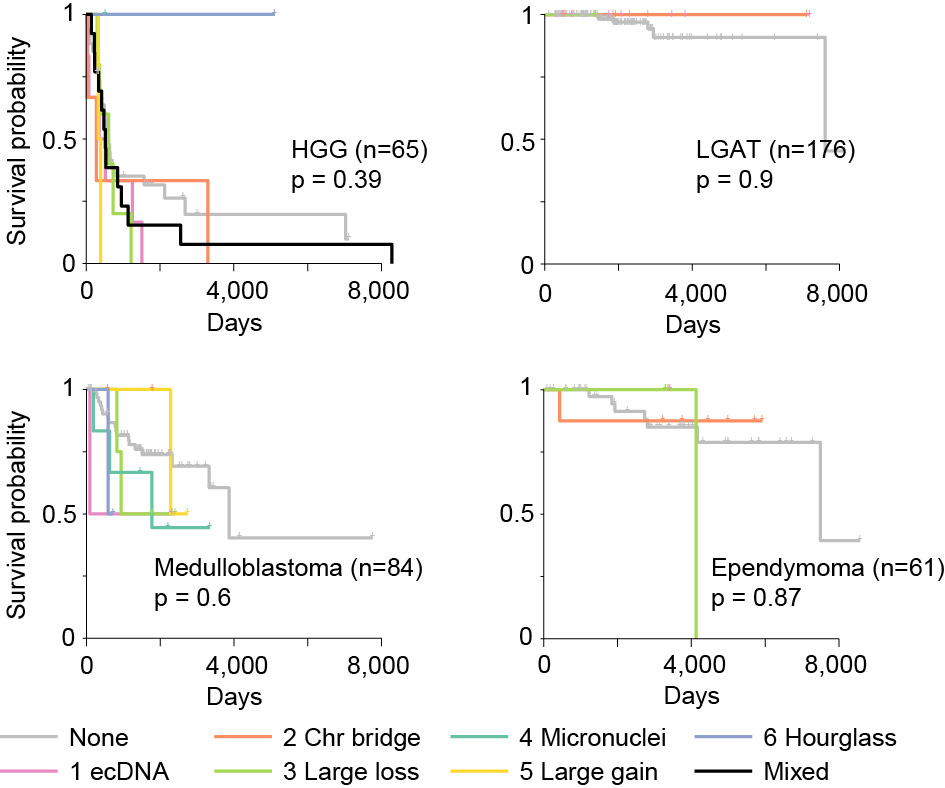

**Figure S4 Kaplan–Meier survival curves for four tumor types.** The samples are colored by the clustered complex SV signatures. Log-rank test was used to calculate p values. The sample sizes are provided in parentheses.
